# A polygenic risk score for coronary heart disease performs well in individuals aged 70 years and older

**DOI:** 10.1101/2021.03.23.21254144

**Authors:** Johannes T. Neumann, Moeen Riaz, Andrew Bakshi, Galina Polekhina, Le T. P. Thao, Mark R. Nelson, Robyn L. Woods, Gad Abraham, Michael Inouye, Christopher M. Reid, Andrew M. Tonkin, John McNeil, Paul Lacaze

## Abstract

**Background:** The use of a polygenic risk score (PRS) to predict coronary heart disease (CHD) events has been demonstrated in the general adult population. However, whether predictive performance extends to older individuals is unclear.

**Aim:** To evaluate the predictive value of a PRS for incident CHD events in a prospective cohort of individuals aged 70 years and older.

**Methods:** We used data from 12,792 genotyped participants of the ASPREE trial, a randomized placebo-controlled trial investigating the effect of daily 100mg aspirin on disability-free survival in healthy older people. Participants had no previous history of diagnosed atherothrombotic cardiovascular events, dementia, or persistent physical disability at enrolment. We calculated a PRS comprising 1.7 million genetic variants (metaGRS). The primary outcome was a composite of incident myocardial infarction or CHD death over 5 years.

**Results:** At baseline, the median population age was 73.9 years and 54.9% were female. In total, 254 incident CHD events occurred. When the PRS was added to conventional risk factors, it was independently associated with CHD (hazard ratio 1.24 [95% confidence interval [CI] 1.08-1.42], p=0.002). The AUC of the conventional model was 70.53 (95%CI 67.00-74.06), and after inclusion of the PRS increased to 71.78 (95%CI 68.32-75.24, p=0.019), demonstrating improved prediction. Reclassification was also improved, as the continuous net reclassification index after adding PRS to the conventional model was 0.25 (95%CI 0.15-0.28).

**Conclusions:** A PRS for CHD performs well in older people, suggesting that the clinical utility of genomic risk prediction for CHD extends to this distinct high-risk subgroup.

## Introduction

An increasing number of recent studies have suggested the potential clinical utility of using a

polygenic risk score (PRS) to improve the prediction of coronary heart disease (CHD) events.^1-8^ It is now well established that individuals in the general population with a high genetic risk score will have higher risk for CHD events, compared to those with a low score.^3^ Furthermore, the addition of a PRS has been shown to significantly improve CHD risk prediction when added to a risk model comprised of conventional risk factors.^4^ PRS performance for CHD risk prediction has recently been validated in more ethnically diverse populations^5, 6^ and populations of European-descent, where improved CHD risk prediction has been shown.^7-9^

The use of genomics in CHD risk prediction has important clinical implications, given the burden of CHD remains high in most countries, despite significant improvements in prevention and treatment. Improved approaches to risk prediction and early intervention may help to address the burden, and genomics presents a new opportunity. However, prior studies investigating a genomic risk scores for CHD risk prediction have mostly included individuals with a mean age ranging from 50 to 60 years or younger. The use of PRS as a risk factor for CHD has not previously been investigated in older individuals specifically, who are themselves a distinct high-risk population.

In addition to the potential differences in PRS performance, older individuals may also require customized CHD risk prediction models with regards to conventional clinical risk factors.^10^ Prediction models used to estimate the risk of future CHD events are usually derived from younger populations and based on conventional risk factors such as blood pressure, diabetes, smoking or blood lipids.^11, 12^ These risk models do not fully explain individual risk in older people, and may require calibration. Here, we sought to investigate the prognostic value of a PRS for CHD in a population of older individuals without a history of CHD events, when added to a conventional risk factor model which we constructed. The objective of our study was to determine whether the potential clinical utility of a PRS for CHD would extend to older individuals aged 70 years and older.

## Methods

### Study design and population

The genotyped population was comprised of participants of the ASPirin in Reducing Events in the Elderly (ASPREE) trial. Study design^13^ and trial results^14, 15^ have been published previously. ASPREE was a randomized double-blind placebo-controlled clinical trial investigating the effect of daily 100mg aspirin on disability-free survival over a median 4.6-years (interquartile range 2.1 years) of follow-up. The trial recruited 19,114 individuals aged ≥70 years (≥ 65 years for US minorities), who had no prior cardiovascular events, and were free from dementia or physical disability at enrolment. Participants had no previous diagnosis of myocardial infarction; heart failure; angina pectoris; stroke; diagnosis of atrial fibrillation; or systolic blood pressure ≥180mmHg. Genetic analyses included 12,792 participants of European descent who provided samples and informed consent (Figure S1). The study was approved by local Ethics Committees and registered on Clinicaltrials.gov (NCT01038583).

### Endpoint

The primary endpoint for this secondary analysis was incident CHD, defined as a composite of incident myocardial infarction or CHD death. CHD death included deaths coded as related to fatal myocardial infarction, sudden cardiac death, rapid cardiac death, or other coronary death. All events were assessed by blinded Adjudication Committees, as described previously^14^.

### Risk model, genotyping and polygenic risk score

The conventional risk model included age, sex, smoking status (current versus former/never), systolic blood pressure, non-high-density-lipoprotein (HDL)-cholesterol, HDL-cholesterol, diabetes and serum creatinine. Selection of variables was based on prior risk models.^12^ Serum creatinine was included based on prior evidence from studies of CHD risk in older individuals.^16, 17^ Aspirin treatment had no effect on CHD risk in the ASPREE population, and was therefore not included in the model (Supplementary results).

Genotyping was performed on 14,052 DNA samples from ASPREE participants using the Axiom 2.0 Precision Medicine Diversity Research Array (Thermo Fisher Scientific, CA, USA) following standard protocols. Variant calling used a custom pipeline aligned to human reference genome hg38. Samples from 12,792 participants passed the following filters: unrelated; Non-Finnish European genetic descent; minimum age at randomization 70 years; and self-reported white racial ancestry. To define genetic descent, we performed principal component analysis (PCA) using the 1000 Genomes reference population and excluded ASPREE samples that did not overlap with the Non-Finnish European 1000 Genomes cluster (Supplementary material, Figure S2).^18^ Imputation was performed using the haplotype reference consortium, European samples (University of Michigan imputation server).^19^ Post-imputation QC removed variants with low imputation quality scores (r^2^<0.3).

We calculated PRS in ASPREE using the metaGRS for CAD^4^ consisting of 1.7 million genetic variants downloaded from the Polygenic Score Catalog.^20^ In the ASPREE data, 1,745,180 (99.6%) of metaGRS SNPs were present (6,140 and 17 SNPs were removed due to variant ID and allele code mismatch, respectively). Plink version 1.9 was used to calculate the weighted sum for effect size of the number of risk alleles for each variant.^21^

### Statistical analyses

Participants with available PRS were included. For continuous variables, the mean and SD are reported. For binary variables, absolute and relative frequencies are provided. Correlation of continuous variables was assessed by Spearman correlation coefficients visualized in a correlation matrix using the package “corrplot”. A multivariable Cox proportional hazards regression model including only predictors from the conventional model was used to evaluate the risk of incident CHD events within 5 years. Continuous variables were used as linear predictors.

The model was re-evaluated after adding the continuous PRS distribution per one SD change, and then by adding PRS divided into tertiles, using the lowest tertile as the reference group, compared with the second and third (higher risk) tertiles. Sensitivity analyses were performed after adding use of antihypertensive drugs, statins and genetic ethnicity PCAs to the multivariable model (Supplementary material). Kaplan-Meier estimates for the incidence of CHD events within 5 years were calculated using the “survival” package and stratified by PRS tertiles.

The area under the curve (AUC) was calculated for each predictor, for the conventional model and after addition of the continuous PRS using time-dependent receiver-operating-characteristics. The analyses were repeated for subgroups according to sex and PRS tertiles (Supplementary material). Reclassification analyses were performed to assess the change in risk after adding the PRS to the conventional model. Time-to-event continuous and categorical net reclassification improvement (NRI) was calculated using the “nricens” package. The risk categories or the categorical NRI were chosen based on the observed risk within the ASPREE cohort and were set to <1.5%, 1.5 to 2.49% and ≥2.5%. Interaction effects between sex and model covariables were examined. All analyses were performed using R version 3.6.1.^22^

## Results

### Baseline characteristics

The median age of the 12,792 participants was 73.9 years (interquartile range 71.7, 77.3, Table 1); 7,027 (54.9%) were female, 391 (3.1%) were current smokers and 1,186 (9.3%) had diabetes. The PRS showed a normal distribution (Figure S3) and the mean value was -1.16 (SD 0.45). There was no relevant correlation of the PRS with other continuous variables within the data set (Figure S4). During follow-up, 254 (2.0%) of subjects had incident CHD events (169 in males, 85 in females). This included 226 incident cases of myocardial infarction and 50 cases of CHD death.

**Table 1:**
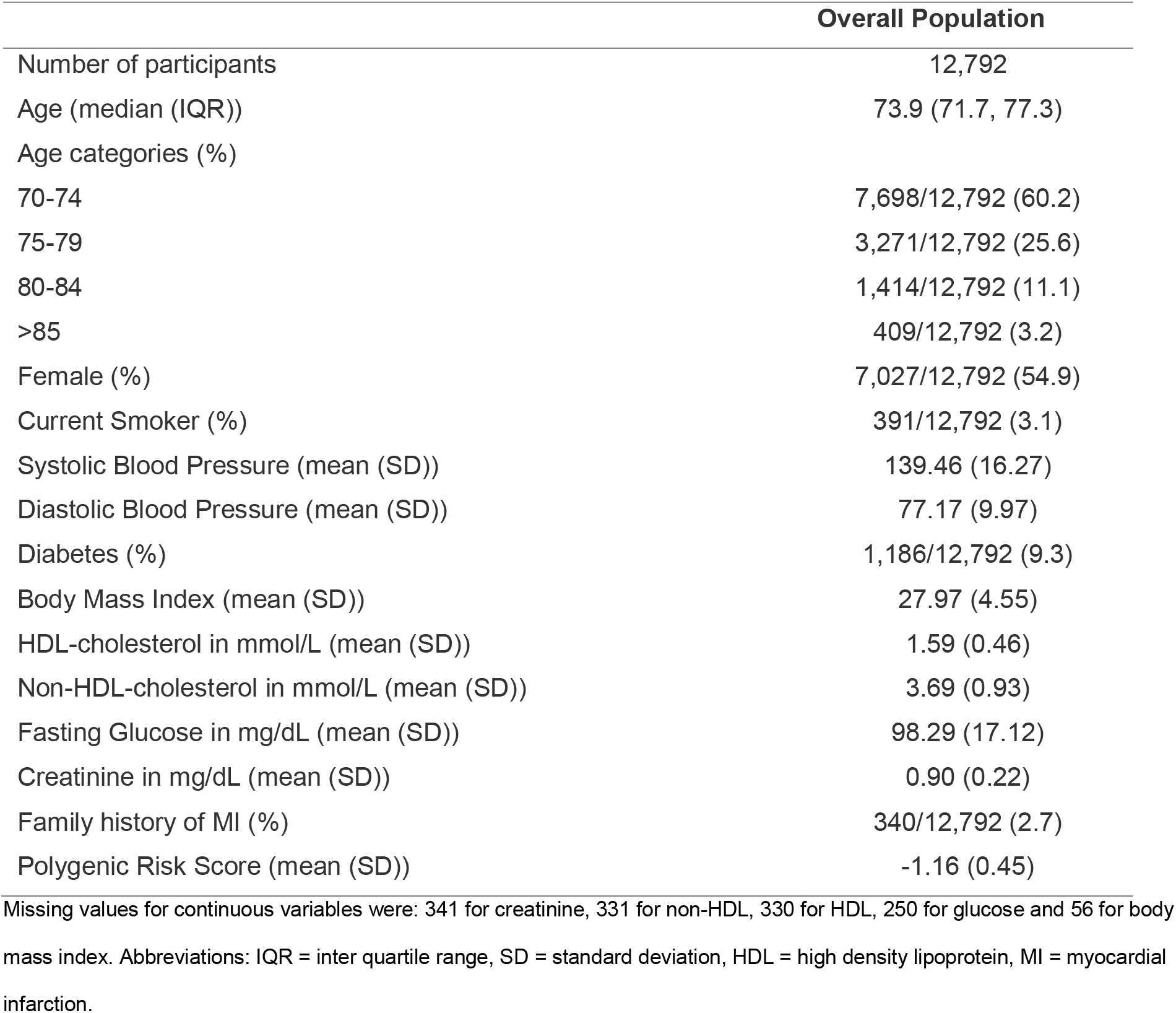
Baseline characteristics

|  | Overall Population |
| --- | --- |
| Number of participants | 12,792 |
| Age (median (IQR)) | 73.9 (71.7, 77.3) |
| Age categories (%) |  |
| 70-74 | 7,698/12,792 (60.2) |
| 75-79 | 3,271/12,792 (25.6) |
| 80-84 | 1,414/12,792 (11.1) |
| >85 | 409/12,792 (3.2) |
| Female (%) | 7,027/12,792 (54.9) |
| Current Smoker (%) | 391/12,792 (3.1) |
| Systolic Blood Pressure (mean (SD)) | 139.46 (16.27) |
| Diastolic Blood Pressure (mean (SD)) | 77.17 (9.97) |
| Diabetes (%) | 1,186/12,792 (9.3) |
| Body Mass Index (mean (SD)) | 27.97 (4.55) |
| HDL-cholesterol in mmol/L (mean (SD)) | 1.59 (0.46) |
| Non-HDL-cholesterol in mmol/L (mean (SD)) | 3.69 (0.93) |
| Fasting Glucose in mg/dL (mean (SD)) | 98.29 (17.12) |
| Creatinine in mg/dL (mean (SD)) | 0.90 (0.22) |
| Family history of MI (%) | 340/12,792 (2.7) |
| Polygenic Risk Score (mean (SD)) | -1.16 (0.45) |

Missing values for continuous variables were: 341 for creatinine, 331 for non-HDL, 330 for HDL, 250 for glucose and 56 for body mass index. Abbreviations: IQR = inter quartile range, SD = standard deviation, HDL = high density lipoprotein, MI = myocardial infarction.

### PRS for risk prediction

In the conventional model, all variables except systolic blood pressure and diabetes were independent predictors of CHD events (Table 2). When the PRS was added as a continuous variable to the model, it was found to be an independent predictor of outcome (Hazard ratio [HR] 1.24 [95% Confidence Interval [CI] 1.08-1.42], p=0.002). Using PRS tertiles as a predictor, CHD risk increased as the PRS category increased from the first to third tertile. When compared to the first PRS tertile (low risk group) the second tertile had a HR for CHD risk of 110 1.48 (95%CI 1.04-2.09, p=0.029) and the third PRS tertile had a HR of 1.64 (95%CI 1.16-2.33, p=0.005). Kaplan-Meier curves illustrated that individuals in the higher and middle PRS tertiles had a higher incidence of CHD events compared with lower PRS tertile (p=0.02, Figure 1).

**Table 2:** Hazard ratios for the conventional model, conventional model + continuous PRS and conventional model + categorical PRS

|  | Conventional model |  |  | Conventional model +<br>continuous PRS |  |  | Conventional model +<br>categorical PRS |  |  |
| --- | --- | --- | --- | --- | --- | --- | --- | --- | --- |
|  | HR | 95%CI | p-value | HR | 95%CI | p-value | HR | 95%CI | p-value |
| Age | 1.09 | (1.06-1.12) | <0.001 | 1.09 | (1.06-1.12) | <0.001 | 1.09 | (1.06-1.12) | <0.001 |
| Female Sex | 0.48 | (0.34-0.67) | <0.001 | 0.46 | (0.33-0.65) | <0.001 | 0.47 | (0.34-0.66) | <0.001 |
| Current Smoking | 2.00 | (1.09-3.68) | 0.025 | 2.02 | (1.10-3.71) | 0.024 | 2.01 | (1.09-3.69) | 0.025 |
| SBP per 10<br>mmHg increase | 1.04 | (0.96-1.13) | 0.34 | 1.04 | (0.96-1.13) | 0.37 | 1.04 | (0.96-1.13) | 0.33 |
| Non-HDL-cholesterol | 1.35 | (1.17-1.56) | <0.001 | 1.35 | (1.17-1.55) | <0.001 | 1.34 | (1.17-1.55) | <0.001 |
| HDL-cholesterol | 0.65 | (0.44-0.95) | 0.026 | 0.65 | (0.44-0.95) | 0.027 | 0.64 | (0.44-0.94) | 0.024 |
| Diabetes | 0.82 | (0.49-1.38) | 0.45 | 0.81 | (0.48-1.36) | 0.42 | 0.81 | (0.48-1.36) | 0.42 |
| Creatinine | 1.83 | (1.03-3.26) | 0.040 | 1.81 | (1.01-3.23) | 0.045 | 1.82 | (1.02-3.24) | 0.043 |
| PRS (continuous<br>per SD) |  |  |  | 1.24 | (1.08-1.42) | 0.002 |  |  |  |
| PRS 1st Tertile |  |  |  |  |  |  | 1.00 | Reference |  |
| PRS 2nd Tertile |  |  |  |  |  |  | 1.48 | (1.04-2.09) | 0.029 |
| PRS 3rd Tertile |  |  |  |  |  |  | 1.64 | (1.16-2.33) | 0.005 |

Abbreviations: SBP = systolic blood pressure, HDL = high density lipoprotein, PRS = polygenic risk score, HR = hazard ratio, CI = confidence interval.

**Figure 1:**
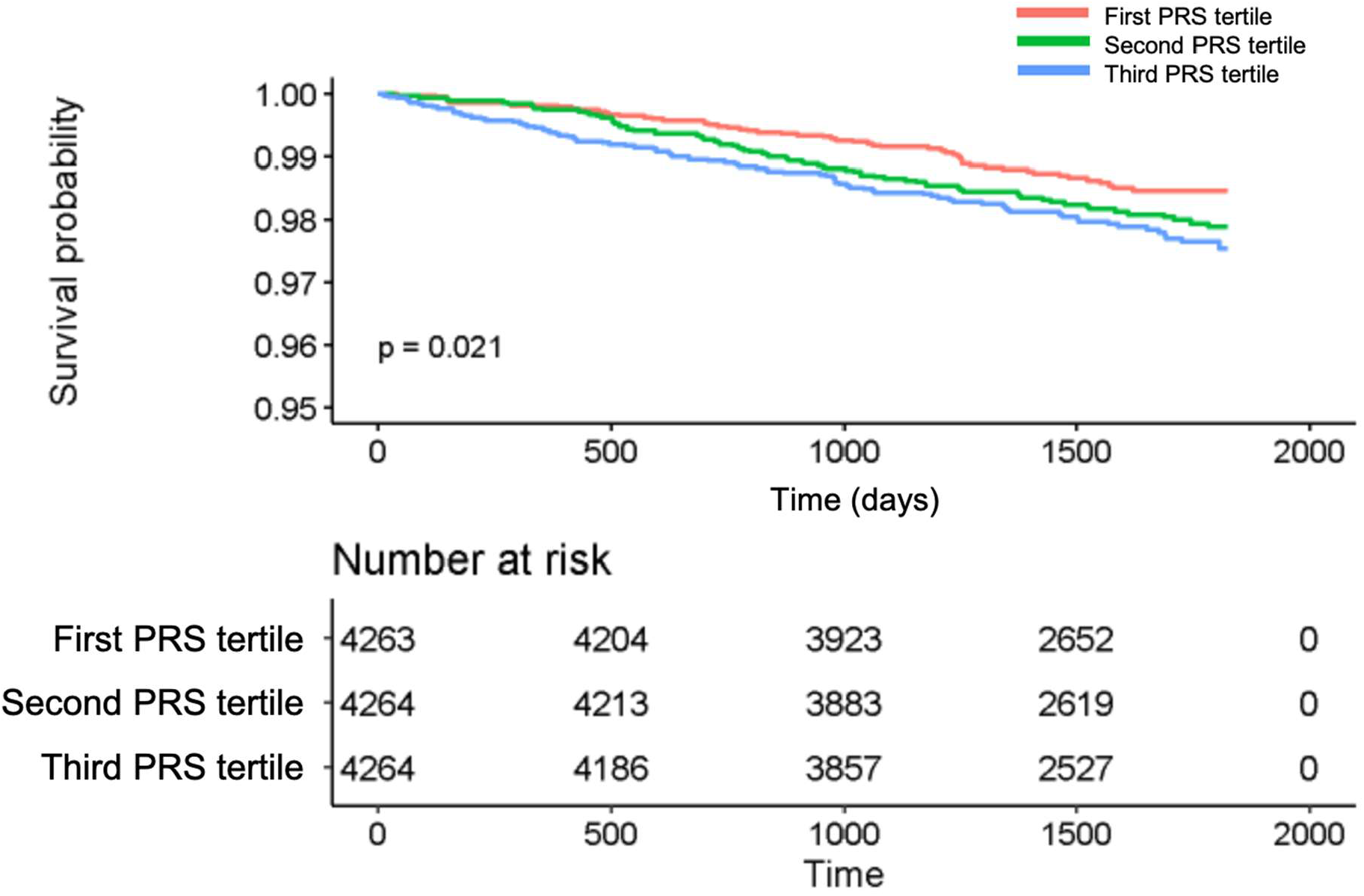
Kaplan-Meier curve for CHD events according to PRS tertiles The figure provides the probability of a CHD event according to tertiles of the PRS, based on Kaplan-Meier estimates, and the individuals at risk.

Evaluation of each predictor using receiver-operating-characteristics showed that sex (AUC 62.88%, 95%CI 59.58-66.17), HDL-cholesterol (AUC 61.56%, 95%CI 57.51-65.61), serum creatinine (AUC 61.39%, 95%CI 57.53-65.24) and age (AUC 57.50%, 95%CI 52.98-62.05) were the strongest predictors of incident CHD events (Figure 2). The PRS alone resulted in an AUC of 55.72% (95%CI 51.74-59.72). The AUC for the conventional model was 70.53% (95%CI 67.00-74.06) and significantly improved to 71.78% (95%CI 68.32-75.24) after adding the PRS as a continuous variable (p=0.019, Table 3, Figure S5). The calibration plot showed a good agreement between predicted and observed CHD events (Figure S6).

**Table 3:** Categorical net reclassification improvement table a<u>fter adding PRS to the conventional model to predict the risk of a 5-year CHD event.</u>

|  |  | Standard Model + Polygenic Risk Score |  |  |  |
| --- | --- | --- | --- | --- | --- |
|  | Standard Model | < 1.5% | 1.5 to 2.49% | ≥ 2.5% | Total No. (%) of participants |
| CHD events | < 1.5% | 37 | 9 | 0 | 46 (22) |
|  | 1.5 to 2.49% | 6 | 35 | 12 | 53 (25) |
|  | ≥ 2.5% | 0 | 8 | 103 | 111 (53) |
|  | Total No. (%) of participants | 43 (20) | 52 (25) | 115 (55) | 210 (100) |
| CHD non-events | < 1.5% | 2248 | 157 | 2 | 2407 (49) |
|  | 1.5 to 2.49% | 204 | 854 | 149 | 1207 (25) |
|  | ≥ 2.5% | 1 | 187 | 1114 | 1302 (26) |
|  | Total No. (%) of participants | 2453 (50) | 1198 (24) | 1265 (26) | 4916 (100) |

**Figure 2:**
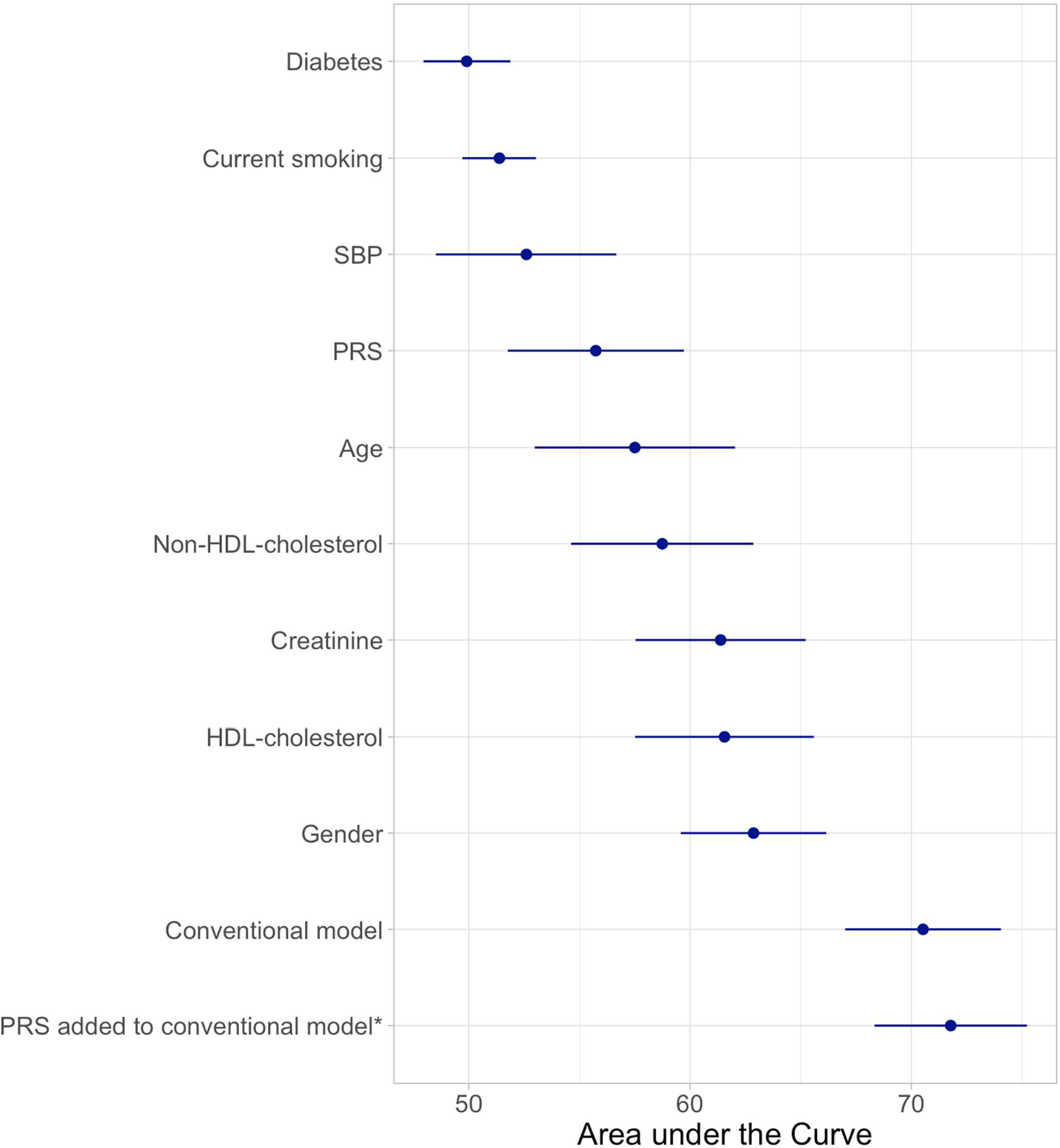
AUC for each predictor, the conventional model and the PRS added to the conventional model *p-value compared the Conventional Model = 0.01899934. Abbreviations: SBP = systolic blood pressure, HDL = high density lipoprotein, PRS = polygenic risk score, AUC = area under the curve, CI = confidence interval.

### Reclassification

In reclassification analyses, the continuous NRI was 0.25 (95%CI 0.15-0.28), when the PRS was added to the conventional model (Table S1). More individuals were to a higher risk category (NRI+ 0.16, 95%CI 0.08-9.20), than downwards (NRI-0.09, 95%CI 0.04-0.10). For measurement of the categorical NRI, CHD risk categories of <1.5%, <2.5% and ≥2.5% were chosen based on the observed risk within ASPREE (Table S1, Table 3). Here, addition of the PRS to the conventional model resulted in a categorical reclassification of 0.063 (95%CI 0.001-0.129), with an upwards classification of 0.044 (95%CI of -0.007-0.105) and a downwards classification of 0.019 (95%CI 0.003-0.032).

### Subgroup analyses

When comparing males and females, we only observed minor differences in baseline characteristics (Table S2). Adding the continuous PRS to the conventional model, it was an independent predictor in males, but not in females (males HR 1.27 [95%CI 1.08-1.50], p=0.005 versus females HR 1.18 [95%CI 0.92-1.49], p=0.19, Table S4+5). The same finding was observed when assessing the categorical PRS. The conventional model resulted in a lower AUC in males compared to females (males AUC 66.58%, females AUC 70.07%), but the incremental value of adding the PRS to the conventional model was greater in males compared with females (males AUC 68.18%, females AUC 71.00%, Table S6).

In subgroup analyses by PRS tertile, baseline characteristics were similar for participants within the highest compared to the lowest PRS tertile (Table S3). The conventional model resulted in a lower AUC in individuals from the highest, compared to individuals from the lowest PRS tertile (highest tertile AUC 73.21%, lowest tertile AUC 76.62%), but the incremental value of addition of the PRS to the conventional model was similar in both groups (Table S7).

Results of sensitivity analyses after adding use of antihypertensive drugs, statins and genetic ethnicity PCAs to the model are reported in the supplementary results (Tables S8+9). Interaction effects between sex and model covariables were examined, but no interaction between sex and PRS was found (HR 0.93, 95%CI 0.69-1.24, p=0.60; Table S10).

## Discussion

In this study, we evaluated the prognostic value of a previously derived polygenic risk score (metaGRS) to predict future CHD events in a population of healthy older individuals from the ASPREE trial. We were able to demonstrate robust PRS performance in this older population and can confirm that addition of the PRS to a conventional cardiovascular risk model improved risk prediction (Figure 3). Our study suggests that the potential clinical utility of a PRS for CHD risk prediction extends to older individuals aged 70 years and older, who comprise an important high-risk group. Our study also represents an independent validation of a PRS recently derived from the UK Biobank, in a well-characterized older population. Our findings add further support to the growing body of evidence that supports the use of genetic information to improve CHD risk prediction, and our results indicate that PRS predictive value extends to older individuals.

**Figure 3:**
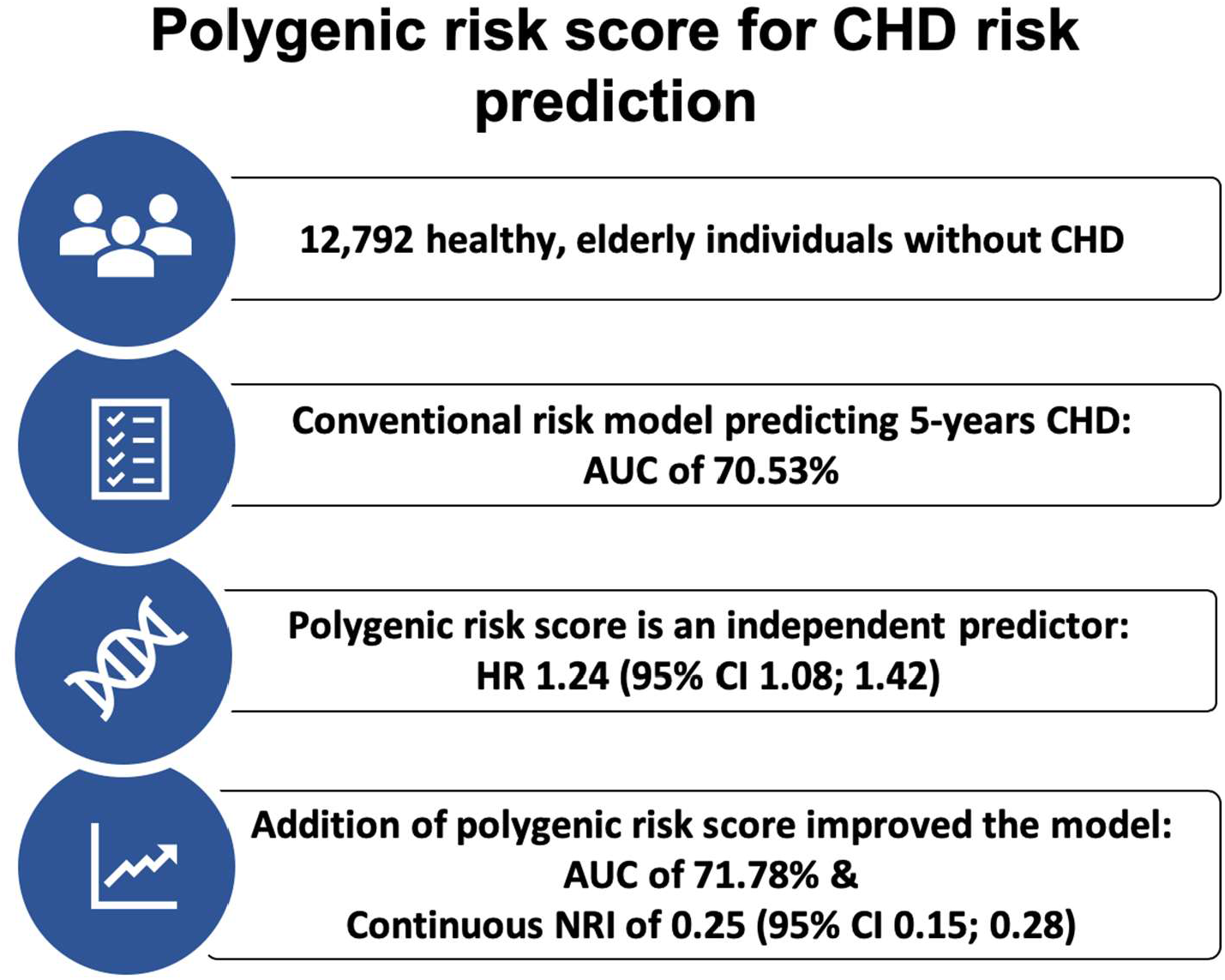
Central figure summarizing the main study findings We evaluated the prognostic accuracy of a previously derived polygenic risk score (metaGRS) to predict 5 years CHD events in a population of healthy elderly individuals. Abbreviations: CHD = coronary heart disease, AUC = area under the curve, HR = hazard ratio, CI = confidence interval.

The metaGRS used in our study was derived from the UK Biobank population of around 500,000 British individuals, with mean age 56.5 years. The ASPREE population differs in several aspects. Firstly, and most notably, the median age of ASPREE participants at enrolment was 73.9 years, nearly 20 years older than the UK Biobank. Second, ASPREE is a highly ascertained clinical trial population, in which participants met strict inclusion criteria, with no history of CHD events at enrolment. Third, major CHD events in ASPREE were adjudicated as part of a randomized trial but did not include coronary revascularization. Given these important differences, it is noteworthy that the metaGRS still performed in a robust manner in the older ASPREE population. Similar to previous studies^5, 6^, our findings demonstrate a polygenic model derived from the UK Biobank generalizes well to other cohorts of European descent.

Cardiovascular disease accounts for a large proportion of deaths in older people. Accurate identification of older individuals at increased risk for CHD is therefore clinically important, particularly those not identified as high-risk by conventional risk factors. Due to a lack of evidence in individuals aged 70 years and older, the value of adding genetic information for CHD risk prediction in older people has not previously been tested robustly. Our study provides the first evidence of its kind to suggest the predictive value and potential clinical utility of a PRS for CHD extends to individuals aged 70 years and older, with comparable predictive performance compared with younger population-based cohorts (refs). Importantly, we found that the PRS alone (considered independently as a CHD risk factor) had similar discriminative power compared to conventional cardiovascular risk factors used in routine practice. However, in our analyses the AUC of sex, HDL-cholesterol, creatinine, non-HDL-cholesterol and age were stronger discriminators, than the PRS alone. This empathizes their role as predictors in an older population, alongside a genetic risk score. Nevertheless, it is noteworthy that the PRS was found to predict CHD events independently of conventional risk factors, not showing correlation with the nine conventional risk factors examined (Figure S4). These unique properties of the genetic risk score (i.e. relatively strong predictive performance and independent effect) help demonstrate its future clinical potential for CHD risk prediction.

Currently, the availability of PRS as a clinical tool for CHD prediction at large remains limited, with unresolved questions related to cost-effectiveness and implementation. Furthermore, some recent studies have provided conflicting results regarding the incremental value of adding genetic information to conventional CHD risk factors in younger populations.^7, 8^ In the future, individual genotyping will become more widely available and at lower cost, potentially facilitating improved CHD event prediction and risk stratification. Here we show that genetic risk is still highly relevant at older ages, and that a PRS for CHD still performs will have may potential clinical utility for preventive strategies in older people. However, further studies of more phenotypically and ethnically diverse elderly populations are required.

Specific findings of our study warrant further discussion. First, we did not find diabetes to be an independent predictor for CHD events, despite 9.3% of ASPREE participants having diabetes at baseline. Other studies have reported the relevance of diabetes regarding CHD risk in the elderly.^10^ This observation could be explained by the pre-selection of a healthy ASPREE population, in whom the duration of diabetes might be shorter, compared to the general population. A second notable finding of our study was that results were not confirmed in subgroup analyses for females. This finding was likely due to limited power because the majority of CHD events in ASPREE occurred in males. Further, we found no interaction effect between sex and PRS, and other studies have reported similar performance for CHD polygenic scores in both sexes.^23^

Strengths of our study include a well-characterized, unique study population with incident cardiovascular events clinically adjudicated as part of a randomized trial. No other large clinical trial has recruited this number of healthy older individuals without a prior history of CHD events, with genotyping. All ASPREE participants received medical assessments by general practitioners at enrolment, to confirm eligibility for the trial, and to rule out previous diagnoses of CHD events. This provided confidence that participants were CHD event-free at enrolment, to examine the value of PRS in the context of primary prevention in the elderly. A range of conventional risk factor variables were also available in ASPREE, to examine alongside polygenic risk.

Limitations of our study include a rather short follow-up period (average 4.6 years per participant) contributing to the relatively small number of CHD events. Continued follow-up will provide more power for future analyses. We also acknowledge the healthy-volunteer effect (ascertained bias) of the ASPREE trial population. ASPREE did not collect information related to revascularization, which is an important CHD endpoint used in metaGRS derivation dataset. Our findings may not be generalizable to other ethnicities or more diverse populations.

In conclusion, we report a potential clinical benefit of using a PRS for improved risk prediction of CHD events in older people. Our study provides evidence that use of PRS for CHD prediction is robust across a diverse range of populations and ages, including individuals aged 70 years and older.

## Data Availability

The data and code that support the findings of this study are available from the corresponding author upon reasonable request.

## Acknowledgements

We thank the ASPREE trial staff, participants, and general practitioners, and the Ramaciotti Centre for Genomics.

