## Supplementary Materials for "A polygenic risk score for coronary heart disease performs well in individuals aged 70 years and older"

**Contentsp**


1. Supplementary Material, page 2
2. Supplementary Results, page 2
3. Supplementary References, page 2
4. Supplementary Tables, page 3
5. Supplementary Figures, page 13

1. Supplementary Material

The genetic ethnicity in the ASPREE cohort were determined through principal component analysis (PCA) using The 1000 Genomes Project as a reference population.[1] Directly genotyped data from ASPREE and The 1000 Genomes Project phase 3 (liftover to hg38) were merged and LD pruned ($r^2 < 0.1$) using plink version 1.9[2], followed by using R package SNPrelate.[3] We calculated the Z score for first 2 PCA and excluded samples with ± 2SD (standard deviation) of Z score compared to their respective five major reference population groups from the 1000 Genomes Project that included: Caucasians (Non-Finish Europeans), South Asians, East Asians, African American and Hispanics (Figure S2).

The first PRS tertile ranged from -2.73 to < -1.35, the second PRS tertile ranged from 1-35 to < -0.97 and the third PRS tertile ranged from -0.97 to 0.69.

2. Supplementary Results

When aspirin treatment was added as covariate to the multivariable model, no significant effect on CHD events was found (HR 0.84, 95%CI 0.64-1.10, p=0.20). In sensitivity analyses, including information on intake of antihypertensive drugs or statins, intake of antihypertensives showed to be a predictor for CHD events (HR 1.73, 95%CI 1.29-2.31), but statin intake did not (HR 0.75, 95%CI 0.53-1.07). However, the PRS remained an independent predictor in the model (Table S7). In sensitivity analyses, including information on the ten PCAs, which were used to determine the genetic ethnicity, none of the PCAs was a significant predictor for CHD events and the PRS remained an independent predictor in the model (Table S8). We examined interaction effects between sex and model covariables but found no interaction effect between sex and the CHD PRS (HR 0.93, 95%CI 0.69-1.24, p=0.60; Table S9).

### 4. Supplementary Tables

Table S1: Estimated net reclassification improvement (NRI, continuous and categorical) for risk of 5-year CHD event.

| Continuous NRI | Estimate | 95%CI |
|---|---|---|
| NRI | 0.25 | 0.15-0.28 |
| NRI+ | 0.16 | 0.08-0.20 |
| NRI- | 0.09 | 0.04-0.10 |
| P(Up\|Case) | 0.58 | 0.54-0.60 |
| P(Down\|Case) | 0.42 | 0.40-0.46 |
| P(Down\|Ctrl) | 0.54 | 0.52-0.55 |
| P(Up\|Ctrl) | 0.46 | 0.45-0.48 |
| **Categorical NRI** | | |
| NRI | 0.063 | 0.001-0.129 |
| NRI+ | 0.044 | -0.007-0.105 |
| NRI- | 0.019 | 0.003-0.032 |
| P(Up\|Case) | 0.106 | 0.031-0.154 |
| P(Down\|Case) | 0.062 | 0.027-0.096 |
| P(Down\|Ctrl) | 0.084 | 0.046-0.116 |
| P(Up\|Ctrl) | 0.066 | 0.037-0.090 |

### Table S2: Baseline characteristics for males and females

| | Male | Female | p-value |
|---|---|---|---|
| Number of participants | 5,765 | 7,027 | |
| Age (median (IQR)) | 73.7 (71.6, 77.0) | 74.0 (71.7, 77.6) | 0.001 |
| Age categories (%) | | | |
| 70-74 | 3,567 (61.9) | 4,131 (58.8) | |
| 75-79 | 1,426 (24.7) | 1,845 (26.3) | |
| 80-84 | 598 (10.4) | 816 (11.6) | |
| >85 | 174 (3.0) | 235 (3.3) | |
| Current Smoker (%) | 198 (3.4) | 193 (2.7) | 0.028 |
| Systolic Blood Pressure (mean (SD)) | 141.41 (15.78) | 137.86 (16.49) | <0.001 |
| Diastolic Blood Pressure (mean (SD)) | 78.07 (9.58) | 76.42 (10.21) | <0.001 |
| Diabetes (%) | 646 (11.2) | 540 (7.7) | <0.001 |
| Body Mass Index (mean (SD)) | 27.93 (3.77) | 28.01 (5.10) | 0.333 |
| HDL in mmol/L (mean (SD)) | 1.40 (0.38) | 1.74 (0.46) | <0.001 |
| Non-HDL in mmol/L (mean (SD)) | 3.63 (0.90) | 3.73 (0.96) | <0.001 |
| Glucose in mg/dL (mean (SD)) | 100.82 (17.99) | 96.22 (16.08) | <0.001 |
| Creatinine in mg/dL (mean (SD)) | 1.02 (0.21) | 0.81 (0.17) | <0.001 |
| Familiy history of MI (%) | 126 (2.2) | 214 (3.0) | 0.003 |
| Polygenic Risk Score (mean (SD)) | -1.18 (0.45) | -1.14 (0.45) | <0.001 |

Table S3: Baseline characteristics according to PRS tertiles

| | First tertile | Second tertile | Third tertile | p-value |
|---|---|---|---|---|
| Number of participants | 4,263 | 4,264 | 4,264 | |
| Age (median (IQR)) | 74.1 (71.8, 77.7) | 73.8 (71.7, 77.3) | 73.7 (71.6, 76.9) | <0.001 |
| Age categories (%) | | | | |
| 70-74 | 2,458 (57.7) | 2,589 (60.7) | 2,651 (62.2) | |
| 75-79 | 1,141 (26.8) | 1,073 (25.2) | 1,057 (24.8) | |
| 80-84 | 522 (12.2) | 455 (10.7) | 436 (10.2) | |
| >85 | 142 (3.3) | 147 (3.4) | 120 (2.8) | |
| Female (%) | 2,239 (52.5) | 2,338 (54.8) | 2,450 (57.5) | <0.001 |
| Current Smoker (%) | 127 (3.0) | 132 (3.1) | 132 (3.1) | 0.937 |
| Systolic Blood Pressure (mean (SD)) | 139.51 (16.36) | 139.18 (15.79) | 139.68 (16.65) | 0.348 |
| Diastolic Blood Pressure (mean (SD)) | 77.15 (10.00) | 77.14 (9.83) | 77.21 (10.07) | 0.944 |
| Diabetes (%) | 381 (8.9) | 375 (8.8) | 430 (10.1) | 0.079 |
| Body Mass Index (mean (SD)) | 27.88 (4.57) | 27.96 (4.58) | 28.07 (4.51) | 0.159 |
| HDL in mmol/L (mean (SD)) | 1.58 (0.46) | 1.59 (0.46) | 1.58 (0.45) | 0.369 |
| Non-HDL in mmol/L (mean (SD)) | 3.66 (0.91) | 3.69 (0.95) | 3.72 (0.94) | 0.019 |
| Glucose in mg/dL (mean (SD)) | 98.13 (17.10) | 98.12 (17.09) | 98.64 (17.19) | 0.291 |
| Creatinine in mg/dL (mean (SD)) | 0.91 (0.22) | 0.91 (0.22) | 0.90 (0.21) | 0.113 |
| Family history of MI (%) | 103 (2.4) | 107 (2.5) | 130 (3.0) | 0.146 |
| Polygenic Risk Score (mean (SD)) | -1.65 (0.24) | -1.16 (0.11) | -0.67 (0.24) | <0.001 |

Table S4: Hazard ratios for the conventional model, conventional model + continuous PRS and conventional model + categorical PRS in males

| | Conventional Model | | | Conventional Model + continuous PRS | | | Conventional Model + categorical PRS | | |
|---|---|---|---|---|---|---|---|---|---|
| | **HR** | **95%CI** | **p-value** | **HR** | **95%CI** | **p-value** | **HR** | **95%CI** | **p-value** |
| Age | 1.05 | (1.02-1.09) | 0.004 | 1.06 | (1.02-1.10) | 0.002 | 1.06 | (1.02-1.10) | 0.002 |
| Current Smoking | 1.99 | (0.97-4.07) | 0.06 | 2.03 | (0.99-4.15) | 0.05 | 2.02 | (0.99-4.13) | 0.05 |
| SBP per 10 mmHg increase | 0.98 | (0.89-1.09) | 0.75 | 0.98 | (0.88-1.09) | 0.71 | 0.98 | (0.89-1.09) | 0.75 |
| Non-HDL | 1.58 | (1.33-1.87) | <0.001 | 1.57 | (1.32-1.86) | <0.001 | 1.56 | (1.32-1.85) | <0.001 |
| HDL | 0.52 | (0.31-0.88) | 0.015 | 0.52 | (0.31-0.88) | 0.015 | 0.52 | (0.31-0.87) | 0.013 |
| Diabetes | 0.97 | (0.54-1.74) | 0.92 | 0.95 | (0.53-1.71) | 0.87 | 0.95 | (0.53-1.70) | 0.86 |
| Creatinine | 1.73 | (0.88-3.38) | 0.11 | 1.71 | (0.87-3.36) | 0.12 | 1.72 | (0.88-3.37) | 0.11 |
| PRS (continuous per SD) | | | | 1.27 | (1.08-1.50) | 0.005 | | | |
| PRS 1st Tertile | | | | | | | 1.00 | (Reference) | |
| PRS 2nd Tertile | | | | | | | 1.49 | (0.97-2.29) | 0.07 |
| PRS 3rd Tertile | | | | | | | 1.79 | (1.17-2.72) | 0.007 |

Table S5: Hazard ratios for the conventional model, conventional model + continuous PRS and conventional model + categorical PRS in females

| | Conventional Model | | | Conventional Model + continuous PRS | | | Conventional Model + categorical PRS | | |
|---|---|---|---|---|---|---|---|---|---|
| | **HR** | **95%CI** | **p-value** | **HR** | **95%CI** | **p-value** | **HR** | **95%CI** | **p-value** |
| Age | 1.14 | (1.09-1.20) | <0.001 | 1.15 | (1.10-1.20) | <0.001 | 1.15 | (1.10-1.20) | <0.001 |
| Current Smoking | 2.26 | (0.71-7.24) | 0.17 | 2.20 | (0.69-7.04) | 0.18 | 2.24 | (0.70-7.17) | 0.17 |
| SBP per 10 mmHg increase | 1.14 | (0.99-1.31) | 0.07 | 1.14 | (0.99-1.31) | 0.07 | 1.14 | (0.99-1.31) | 0.06 |
| Non-HDL | 1.00 | (0.78-1.29) | 1.00 | 1.00 | (0.77-1.29) | 0.99 | 1.00 | (0.77-1.29) | 0.99 |
| HDL | 0.77 | (0.44-1.38) | 0.39 | 0.78 | (0.44-1.39) | 0.40 | 0.77 | (0.43-1.37) | 0.38 |
| Diabetes | 0.52 | (0.16-1.68) | 0.27 | 0.51 | (0.16-1.67) | 0.27 | 0.51 | (0.16-1.67) | 0.27 |
| Creatinine | 1.81 | (0.55-5.95) | 0.33 | 1.78 | (0.55-5.83) | 0.34 | 1.78 | (0.55-5.80) | 0.34 |
| PRS (continuous per SD) | | | | 1.18 | (0.92-1.49) | 0.19 | | | |
| PRS 1st Tertile | | | | | | | 1.00 | (Reference) | |
| PRS 2nd Tertile | | | | | | | 1.37 | (0.75-2.52) | 0.31 |
| PRS 3rd Tertile | | | | | | | 1.36 | (0.74-2.49) | 0.32 |

Table S6: AUC for each predictor, the conventional model and the PRS added to the conventional model in males and females

| | Males | | Females | |
|---|---|---|---|---|
| | **AUC** | **95%CI** | **AUC** | **95%CI** |
| Age | 53.41% | (47.75-59.08) | 67.35% | (60.06-74.65) |
| Current Smoking | 51.24% | (49.26-53.22) | 51.37% | (48.35-54.40) |
| Systolic Blood Pressure | 48.15% | (42.96-53.34) | 57.32% | (50.71-63.93) |
| Non-HDL | 63.85% | (58.81-68.89) | 50.79% | (43.58-58.00) |
| HDL | 58.30% | (53.55-63.05) | 50.71% | (43.35-58.08) |
| Diabetes | 49.86% | (47.24-52.49) | 51.02% | (48.42-53.62) |
| Creatinine | 54.99% | (49.66-60.34) | 56.01% | (49.72-62.30) |
| PRS | 57.15% | (52.20-62.11) | 54.98% | (48.21-61.77) |
| Conventional Model | 66.58% | (61.89-71.29) | 70.07% | (63.46-76.69) |
| PRS added to Conventional Model | 68.18% | (63.64-72.73) | 71.00% | (64.45-77.55) |

Table S7: AUC for each predictor, the conventional model and the PRS added to the conventional model in individuals from the lowest and highest PRS tertile

| | Lowest PRS Tertile | | Highest PRS Tertile | |
|---|---|---|---|---|
| | **AUC** | **95%CI** | **AUC** | **95%CI** |
| Age | 65.35% | (56.82-73.88) | 50.62% | (43.02-58.22) |
| Sex | 61.58% | (55.28-67.89) | 64.76% | (59.38-70.15) |
| Current Smoking | 49.99% | (47.72-52.28) | 52.67% | (49.58-55.78) |
| Systolic Blood Pressure | 52.86% | (44.97-60.75) | 52.81% | (46.34-59.29) |
| Non-HDL | 64.85% | (57.54-72.16) | 61.68% | (55.62-67.75) |
| HDL | 54.94% | (47.13-62.76) | 67.20% | (60.82-73.59) |
| Diabetes | 51.94% | (49.67-54.23) | 50.61% | (47.47-53.76) |
| Creatinine | 61.06% | (53.22-68.91) | 59.35% | (53.36-65.34) |
| PRS | 50.37% | (42.60-58.15) | 51.55% | (44.59-58.53) |
| Conventional Model | 76.62% | (70.49-82.76) | 73.21% | (67.39-79.03) |
| PRS added to Conventional Model | 77.15% | (71.07-83.23) | 73.38% | (67.63-79.15) |

Table S8: Sensitivity analyses including information on intake of antihypertensive drugs and statins for calculation of the multivariable Cox regression model.

| | HR | 95%CI | p-value |
|---|---|---|---|
| Age | 1.09 | (1.06-1.12) | <0.001 |
| Female Sex | 0.44 | (0.31-0.61) | <0.001 |
| Current Smoking | 2.05 | (1.12-3.77) | 0.021 |
| SBP per 10 mmHg increase | 1.02 | (0.94-1.11) | 0.57 |
| Non-HDL | 1.34 | (1.15-1.56) | <0.001 |
| HDL | 0.67 | (0.46-0.99) | 0.043 |
| Diabetes | 0.79 | (0.47-1.34) | 0.38 |
| Creatinine | 1.58 | (0.88-2.83) | 0.13 |
| PRS (continuous per SD) | 1.23 | (1.08-1.41) | 0.003 |
| Intake of antihypertensives | 1.73 | (1.29-2.31) | <0.001 |
| Intake of statin | 0.75 | (0.53-1.07) | 0.11 |

Table S9: Sensitivity analyses including information on PCAs for calculation of the multivariable Cox regression model.

| | HR | 95%CI | p-value |
|---|---|---|---|
| Age | 1.09 | (1.06; 1.12) | <0.001 |
| Female Sex | 0.46 | (0.33; 0.65) | <0.001 |
| Current Smoking | 2.02 | (1.10; 3.71) | 0.024 |
| SBP per 10 mmHg increase | 1.04 | (0.96; 1.13) | 0.36 |
| Non-HDL | 1.35 | (1.17; 1.56) | <0.001 |
| HDL | 0.64 | (0.44; 0.95) | 0.025 |
| Diabetes | 0.81 | (0.48; 1.36) | 0.42 |
| Creatinine | 1.88 | (1.05; 3.35) | 0.033 |
| PRS (continuous per SD) | 1.25 | (1.09; 1.43) | 0.002 |
| PCA1 | 0.87 | (0.50; 1.51) | 0.61 |
| PCA2 | 1.00 | (0.66; 1.53) | 0.98 |
| PCA3 | 1.14 | (0.93; 1.40) | 0.19 |
| PCA4 | 1.12 | (0.92; 1.36) | 0.27 |
| PCA5 | 0.98 | (0.93; 1.02) | 0.28 |
| PCA6 | 0.98 | (0.96; 1.01) | 0.24 |
| PCA7 | 1.00 | (0.97; 1.02) | 0.79 |
| PCA8 | 0.99 | (0.93; 1.05) | 0.70 |
| PCA9 | 1.00 | (0.99; 1.02) | 0.56 |
| PCA10 | 1.01 | (0.99; 1.03) | 0.61 |

Table S10: Interaction of sex with model covariables.

| | HR | 95% CI | p-value |
|---|---|---|---|
| Age | 1.06 | (1.02; 1.10) | 0.002 |
| Female Sex | 0.00 | (0.00; 0.06) | 0.002 |
| Current Smoking | 2.02 | (0.99; 4.14) | 0.05 |
| SBP per 10 mmHg increase | 0.98 | (0.88; 1.09) | 0.72 |
| Non-HDL | 1.57 | (1.32; 1.86) | <0.001 |
| HDL | 0.52 | (0.31; 0.88) | 0.015 |
| Diabetes | 0.95 | (0.53; 1.70) | 0.86 |
| Creatinine | 1.71 | (0.87; 3.36) | 0.12 |
| PRS (continuous per SD) | 1.27 | (1.08; 1.50) | 0.005 |
| Female Sex * Age | 1.08 | (1.02; 1.15) | 0.007 |
| Female Sex * Current Smoking | 1.09 | (0.28; 4.27) | 0.90 |
| Female Sex * SBP per 10 mmHg increase | 1.16 | (0.98; 1.38) | 0.09 |
| Female Sex * Non-HDL | 0.64 | (0.47; 0.86) | 0.004 |
| Female Sex * HDL | 1.49 | (0.68; 3.24) | 0.31 |
| Female Sex * Diabetes | 0.54 | (0.15; 2.02) | 0.36 |
| Female Sex * Creatinine | 1.04 | (0.27; 4.08) | 0.95 |
| Female Sex * PRS (continuous per SD) | 0.93 | (0.69; 1.24) | 0.60 |

5. Supplementary Figures

Figure S1: Study flow chart

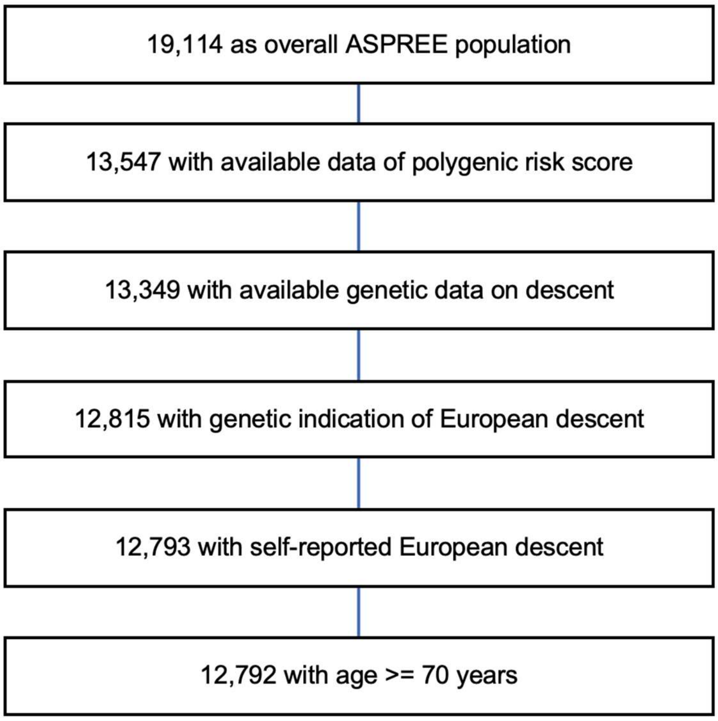

Figure S2: Principal component analysis (PCA) of the ASPREE cohort compared with the 1000 Genome Project. A. Shows PCA plot of all ASPREE participants mapped with 1000 Genome populations. B. Shows PCA plot of ASPREE Europeans samples with 1000 Genome Europeans samples that were included in this study.

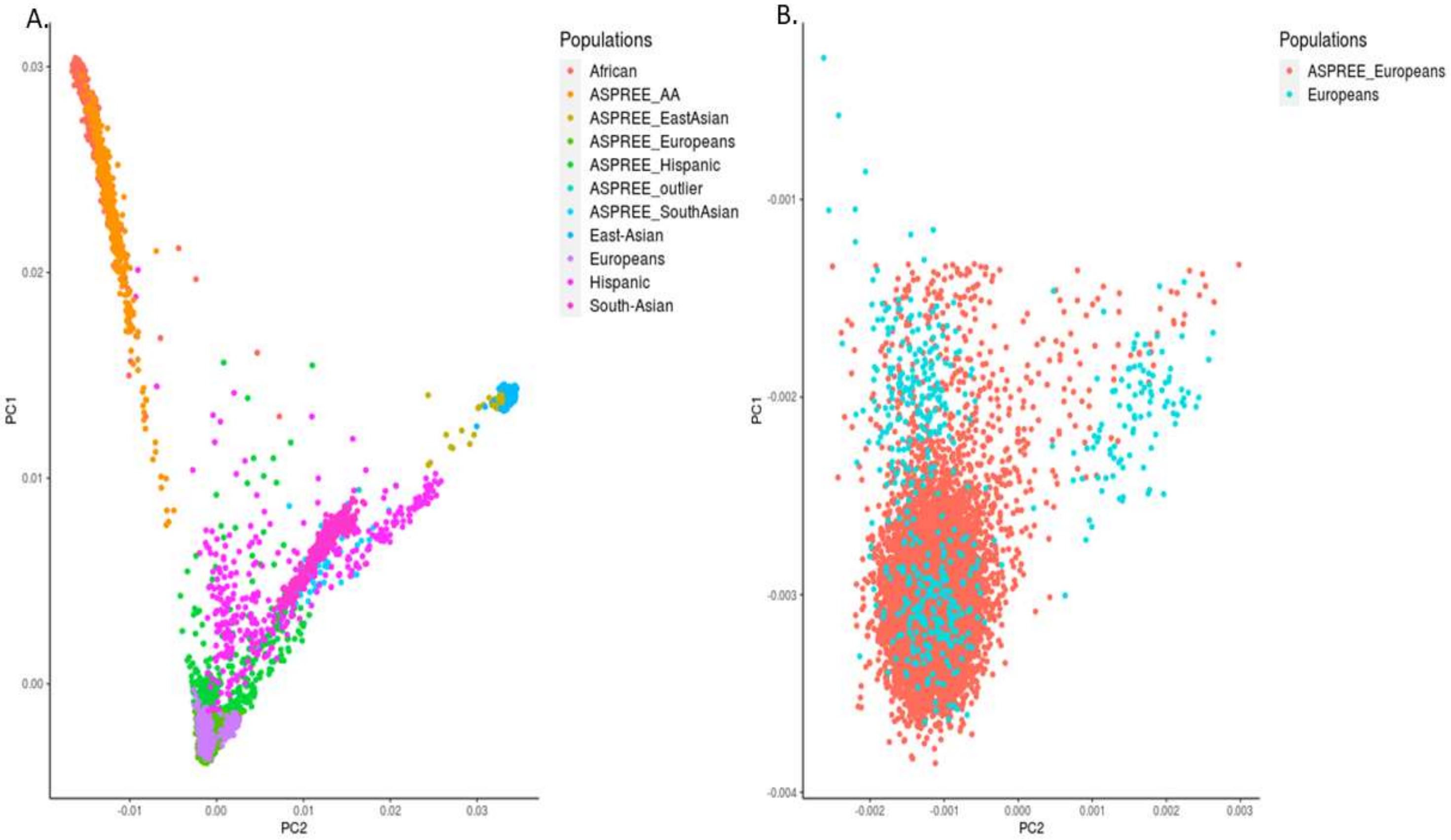


In Figure legend 1000 genome populations are: Europeans, South Asians, East Asians, African American and Hispanics. ASPREE_AA is African American samples.

Figure S3: Distribution of the Polygenic Risk Score in the study population

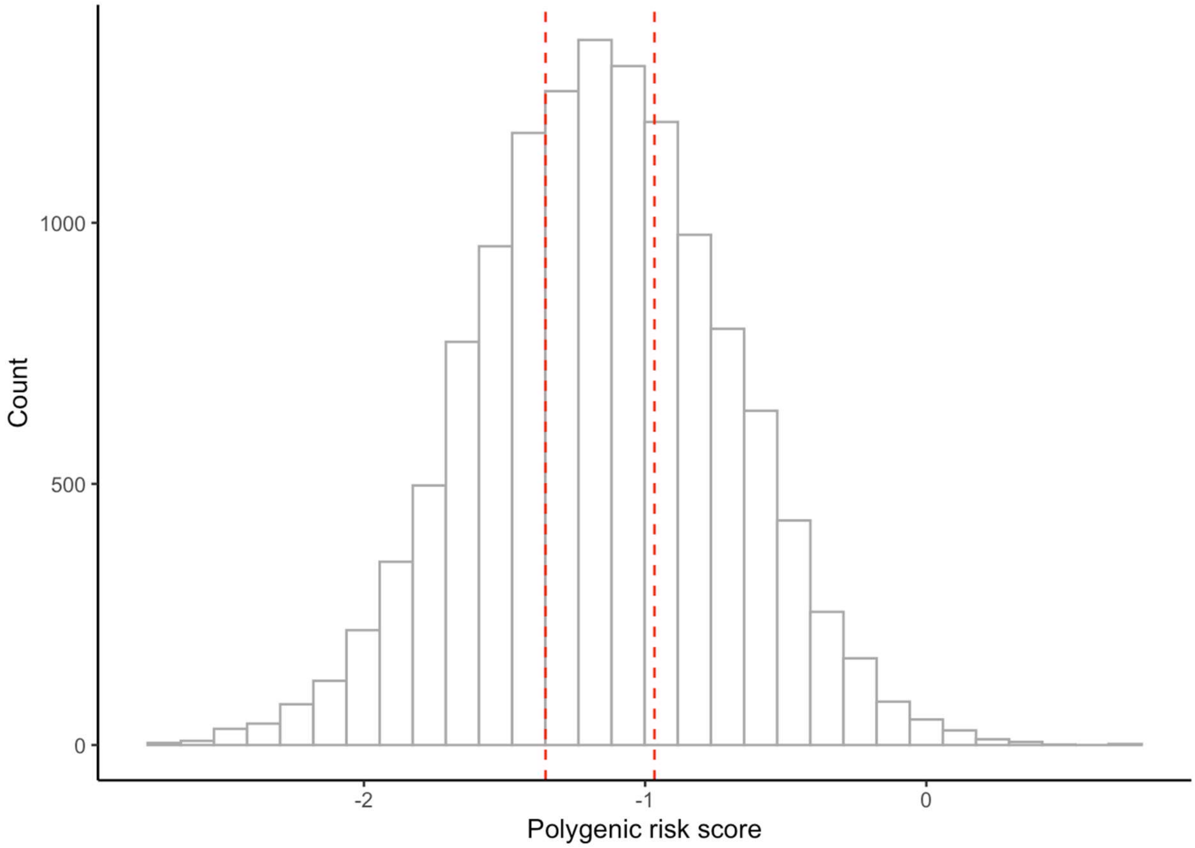


The distribution of the PRS in the study population is provided. The mean value was -1.16 (SD 0.45). The red dashed lines indicate the PRS tertiles. The first tertile ranged from -2.73 to < -1.35, the second tertile ranged from 1-35 to < -0.97 and the third tertile ranged from -0.97 to 0.69.

Figure S4: Correlation matrix of continuous variables including the Polygenic Risk Score

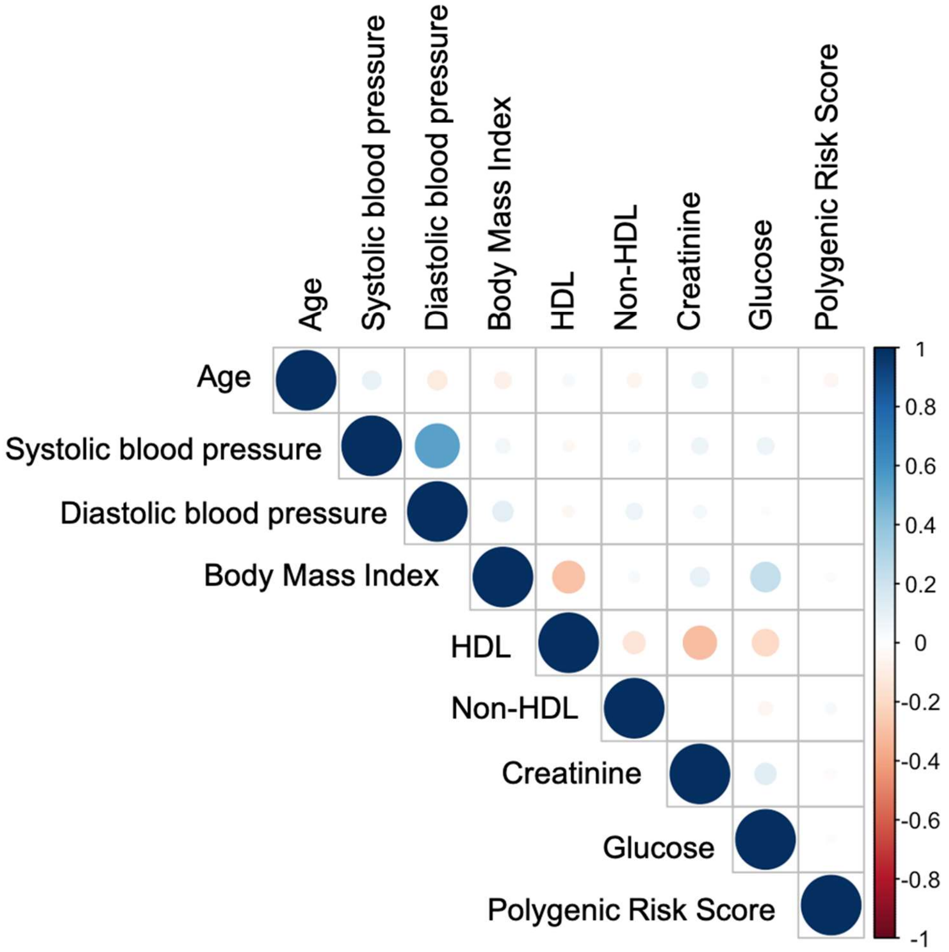


A correlation matrix including only continuous variables is provided. The size and the color of the circles indicate the strength of correlation, ranging from r = -1 (red) to +1 (blue).

Figure S5: AUC for the conventional model and after addition of the PRS to the conventional model

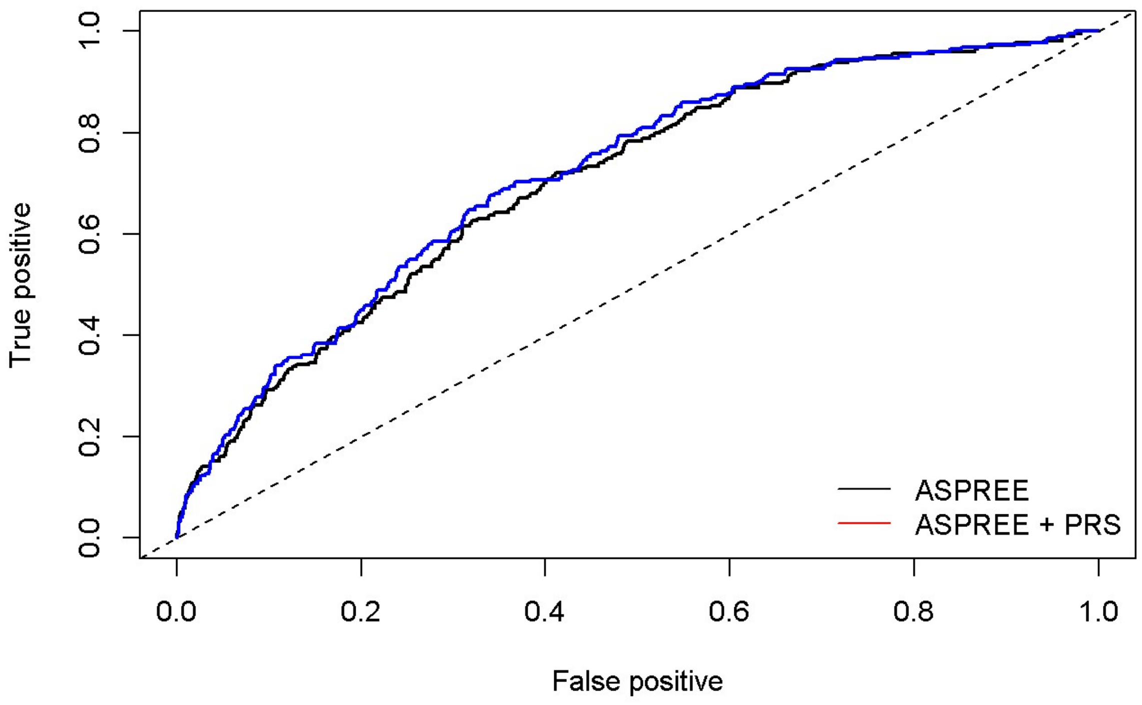


The area under the curve is provided for the conventional model including age, sex, smoking status (current smoking versus former or never smoking), systolic blood pressure, non-HDL, HDL, diabetes and creatinine. In addition, the area under the curve is re-calculated after adding the PRS to the risk model to discriminate between CHD events and non-events.

Figure S6: Calibration of the conventional model + continuous PRS

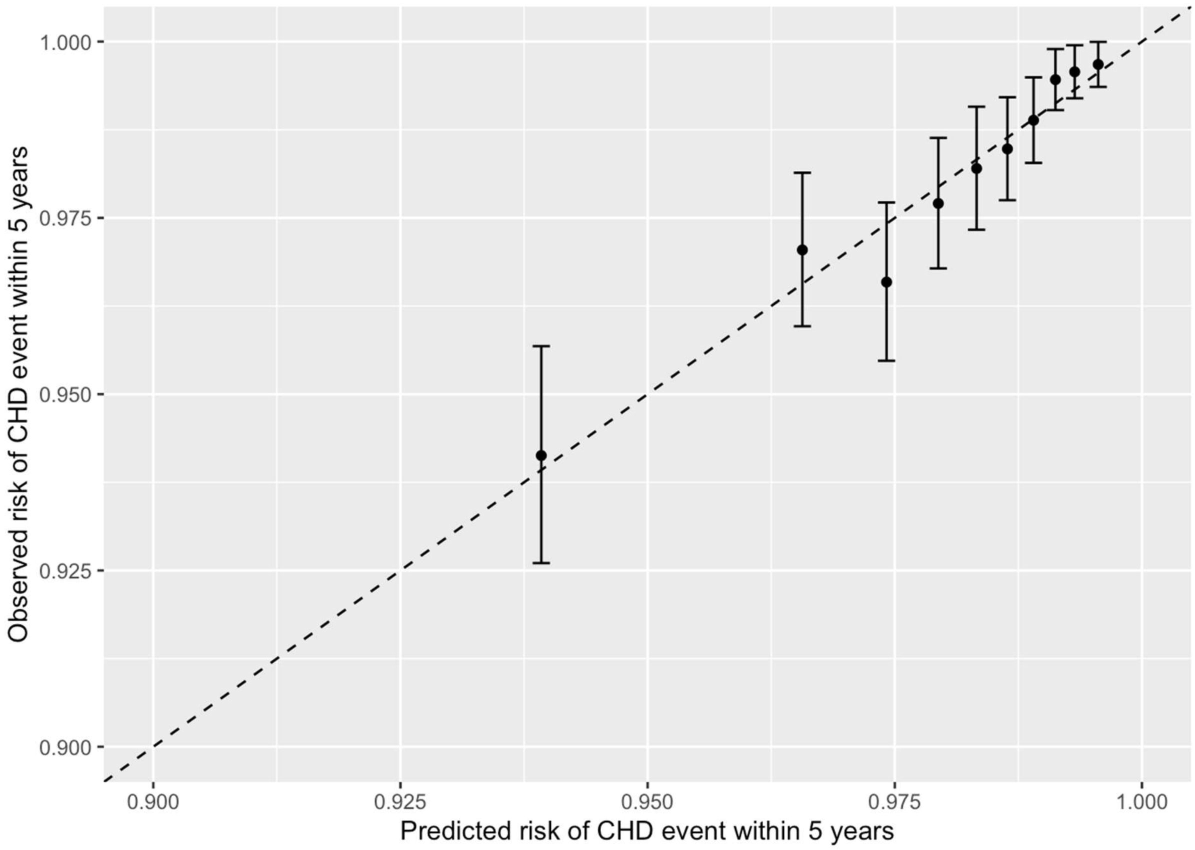


In this figure the observed and predicted risk for CHD events is compared.
